# Developing and Testing an Engineering Framework for Curiosity-Driven and Humble AI in Clinical Decision Support

**DOI:** 10.64898/2026.02.06.26345664

**Authors:** Janan Arslan, Kurt Benke, Sebastián Andrés Cajas Ordoñez, Rowell Castro, Leo Anthony Celi, Gustavo Adolfo Cruz-Suarez, Roben Delos Reyes, Justin Engelmann, Ari Ercole, Almog Hilel, Mahima Kalla, Leo Kinyera, Maximin Lange, Torleif Markussen Lunde, Mackenzie J Meni, Felipe Ocampo Osorio, Anna E Premo, Jana Sedlakova, Pritika Vig

## Abstract

**Background:** We present BODHI (Balanced, Open-minded, Diagnostic, Humble, and Inquisitive), an engineering framework for curiosity-driven and humble clinical decision support AI. Despite growing capabilities, large language models (LLMs) often express inappropriate confidence, conflating statistical pattern recognition with genuine medical understanding. BODHI addresses this through a dual-reflective architecture that: (1) decomposes epistemic uncertainty into task-specific dimensions, and (2) constrains model responses using virtue-based stance rules derived from a Virtue Activation Matrix.

**Methods:** We validate the framework through controlled evaluation on 200 clinical vignettes from HealthBench Hard, assessing GPT-4o-mini and GPT-4.1-mini across 5 random seeds (1,800 total observations). Statistical analysis included bootstrap resampling, paired t-tests, and effect size computation (Supplementary Materials S3)

**Findings:** BODHI significantly improved overall clinical response quality (GPT-4.1-mini: +17.3pp, *p <* 0.0001, Cohen’s *d* = 0.50; GPT-4o-mini: +7.4pp, *p <* 0.0001, Cohen’s *d* = 0.22) while achieving very large effect sizes on curiosity (context-seeking rate: Cohen’s *d* = 16.38 and 19.54) and humility (hedging: *d* = 5.80 for GPT-4.1-mini) metrics. Crucially, 97.3% of GPT-4.1-mini responses and 73.5% of GPT-4o-mini responses included appropriate clarifying questions, compared to 7.8% and 0.0% at baseline, demonstrating the framework’s effectiveness in eliciting information-gathering behavior.

**Interpretation:** These findings suggest LLMs can be reliably constrained to operate within epistemic boundaries when provided with structured uncertainty decomposition and virtue-aligned response rules, offering a pathway toward safer clinical AI deployment.

## 1 Introduction

Medical errors remain a leading cause of death, claiming over 250,000 lives annually in the United States [1]. Artificial Intelligence (AI) systems that express unwarranted confidence can contribute to such errors in clinical settings: experienced radiologists might follow incorrect AI suggestions despite contradictory evidence [2], while ICU clinicians defer to systems even when clinical intuition suggests otherwise [3, 4]. This often happens when presented with high confidence scores [2].

Large language models (LLMs) regularly exhibit overconfidence in clinical reasoning tasks. Recent benchmarking evaluations show that even the most accurate LLMs demonstrate minimal variation in confidence between correct and incorrect answers, limiting their safe use in clinical settings [5]. LLMs show inflexible reasoning and propensity to hallucinate when faced with scenarios requiring deviation from training patterns [6], and exhibit sycophantic behavior (excessively praising or flattering a person in authority, in this case the prompter/user) with up to 100% compliance, even when presented with illogical medical requests [7]. Uncertainty estimation analyses indicate overconfidence despite limited accuracy [6, 7], with confidence scores failing to align with prediction accuracy across models and medical specialties [8].

Several approaches have attempted to address AI overconfidence. Uncertainty quantification methods and conformal prediction can estimate epistemic uncertainty [9–11]. However, these approaches focus on measuring uncertainty rather than modifying behavior when uncertain. Lindenmeyer et al. [9] compared ensemble and Gaussian process approaches for uncertainty estimation in clinical decision support, yet systems still produced confident outputs regardless of estimated uncertainty. Chain-of-thought prompting has improved medical reasoning accuracy and interpretability [12–14], but existing implementations optimize for correctness rather than appropriate epistemic conduct. Fine-tuning approaches like ConfiDx [15] show promise for uncertainty-aware diagnosis but require model modification and focus narrowly on diagnostic tasks.

Previous work has called for epistemic humility as a design principle [16] arguing epistemic humility in AI-assisted pain assessment requires ethics education for developers and clinicians, but stops short of engineering implementations. Madhukar [17] formalizes the ‘AI Socratic Paradox’ where clinical AI must know when to deviate from its own recommendations, highlighting the need for metacognitive self-awareness but not providing concrete solutions. Uncertainty is often quantified without behavioral modification and epistemic virtues discussed without operationalization, or they require model fine-tuning that limits practical deployment. No prompting-based framework has demonstrated empirical effectiveness in constraining LLM behavior to align with epistemic virtues in clinical contexts.

We previously introduced curiosity and humility as essential epistemic virtues for healthcare AI [18–20]. Curiosity represents the drive to reduce uncertainty through targeted inquiry, while humility involves acknowledging limitations and deferring appropriately to human expertise. Building on this conceptual foundation, the present work addresses the critical gap between philosophical proposals and engineering implementation. Our framework for balanced, open-minded, diagnostic, humble, and inquisitive machines (BODHI) fills this gap through a two-pass chain-of-thought protocol. The first pass generates structured uncertainty analysis; the second produces appropriately constrained responses guided by a Virtue Activation Matrix. We validate BODHI on 200 clinical vignettes from HealthBench Hard [21], demonstrating improvements in context-seeking behavior.

## 2 Methods

### 2.1 Framework Architecture

The BODHI framework operates through six integrated steps (Supplementary Materials Figure S1). First, clinical complexity assessment evaluates the incoming query across multiple dimensions including diagnostic ambiguity, urgency indicators, and data completeness. Second, prior confidence evaluation estimates the model’s epistemic state based on training distribution coverage and query specificity. Third, parallel Curiosity and Humility modules process the clinical scenario: the Curiosity module identifies information gaps and generates clarifying questions, while the Humility module assesses confidence boundaries and deferral triggers. Fourth, the Virtue Activation Matrix maps the combined outputs to one of four epistemic stances. Fifth, adaptive system responses are generated according to the selected stance. Sixth, continuous learning mechanisms incorporate clinician feedback to refine threshold calibrations over time.

### 2.2 Virtue Activation Matrix

The Virtue Activation Matrix (Figure 1) maps clinical complexity against model confidence to determine appropriate epistemic stances. Four quadrants organize behavioral responses based on the interplay between uncertainty and stakes.

**Figure 1:**
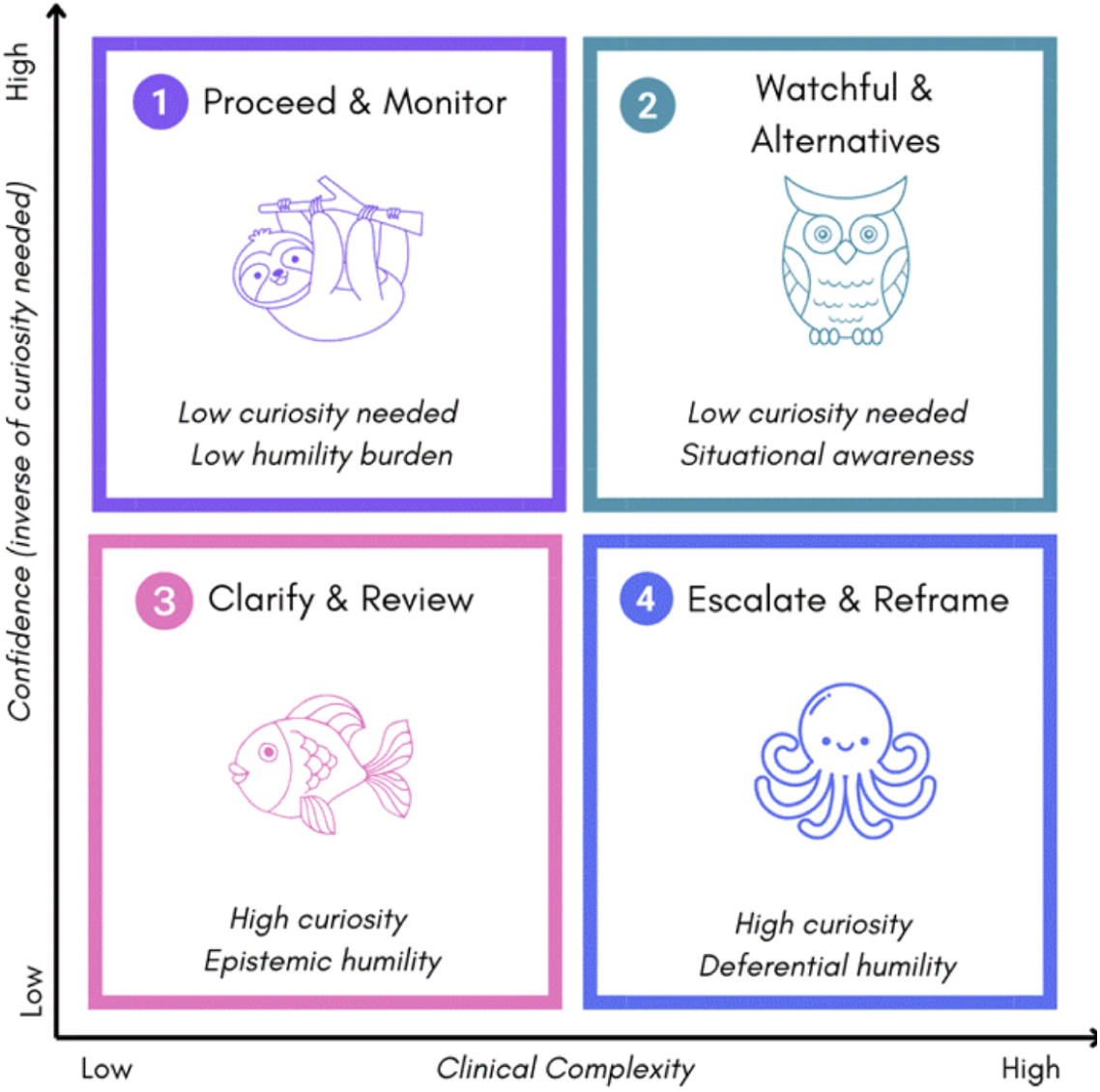
Virtue Activation Matrix mapping clinical complexity against model confidence. Each quadrant represents a distinct epistemic stance with associated curiosity and humility requirements. Q1 (Proceed & Monitor): routine cases with high confidence requiring low curiosity and low humility burden. Q2 (Watchful & Alternatives): complex cases requiring situational awareness. Q3 (Clarify & Review): uncertain cases needing high curiosity and epistemic humility. Q4 (Escalate & Reframe): high-stakes uncertainty demanding high curiosity and deferential humility with explicit escalation to human expertise.

Quadrant 1: Proceed and Monitor applies when confidence is high and complexity is low, requiring minimal curiosity and low humility burden.

Quadrant 2: Watchful and Alternatives activates under high confidence but high complexity, demanding situational awareness and consideration of differential diagnoses.

Quadrant 3: Clarify and Review governs low confidence with low complexity scenarios, triggering high curiosity and epistemic humility through targeted clarifying questions.

Quadrant 4: Escalate and Reframe represents the most demanding stance: low confidence under high complexity requires high curiosity combined with deferential humility, explicitly escalating to human expertise.

The decision logic for stance selection follows a tree of conditional probabilities based on clinical complexity indicators (Supplementary Materials Figure S2). Operational thresholds for confidence and complexity classification are detailed in Supplementary Materials S5.1.

### 2.3 Two-Pass Chain-of-Thought Protocol

The BODHI protocol operationalizes epistemic virtues through structured chain-of-thought scaffolding that separates internal reasoning from external communication.

Pass 1 generates structured analysis with seven mandatory fields: task type classification (Emergency, Technical, Hybrid, or Conversation), audience identification (Patient, Health Professional, or Unclear), primary hypothesis with reasoning, key uncertainties affecting confidence, clarifying questions (1 to 2 required for non-emergency cases), red flags triggering escalation, and safe recommendations appropriate to the uncertainty level. This structured output makes the model’s epistemic state explicit and auditable.

Pass 2 produces the clinician-facing response by conditioning on Pass 1 analysis and applying epistemic constraints. Task routing rules adjust behavior based on context: Emergency Mode prioritizes safety over completeness, Technical Mode reduces hedging for administrative tasks, Hybrid Mode balances clinical reasoning with technical precision, and Conversation Mode (default) applies full epistemic constraints for patient-facing interactions. Cross-cutting constraints enforce specificity (concrete numbers and timeframes when available), active inquiry (converting conditional statements into direct questions), and presentation of alternatives (acknowledging differential diagnoses when confidence is below threshold). Complete protocol specifications including the full prompting templates are provided in Supplementary Materials S2.

### 2.4 Evaluation Design

We evaluated BODHI using HealthBench Hard [21], a benchmark of 200 challenging clinical scenarios requiring diagnostic reasoning, treatment planning, and triage decisions. HealthBench Hard represents a curated subset selected for cases requiring nuanced clinical judgment where baseline models show suboptimal performance. The dataset spans multiple clinical domains including emergency medicine, primary care, and specialty consultations.

Two frontier language models were assessed: GPT-4.1-mini and GPT-4o-mini across 5 random seeds (42, 43, 44, 45, 46) yielding 1000 case-level observations. Each model was evaluated under baseline (standard prompting) and BODHI (two-pass protocol) conditions. This seed-based replication design enables quantification of reproducibility and controls for sampling variance. Model configuration details including temperature (0.7), token limits (4096), and API parameters are provided in Supplementary Materials S3.2, Table S3.

Statistical analysis employed nonparametric bootstrap resampling (10,000 iterations) for confidence intervals, case-level paired t-tests accounting for repeated measures structure, mixed-effects models with random intercepts for seed and case, and Cohen’s *d* with Hedge’s *g* correction for effect sizes (Supplementary Materials S3.4). Primary outcomes included overall HealthBench score (composite rubric aggregating accuracy, safety, and communication), context-seeking rate (proportion of responses containing at least one appropriate clarifying question), hedging frequency (explicit uncertainty acknowledgment), helpful and safe score, emergency referral rate, and communication quality. Complete metric definitions are provided in Supplementary Materials S4.1, Table S4.

## 3 Results

### 3.1 Primary Outcomes

BODHI produced significant improvements across both models (Table 1). For GPT-4.1-mini, overall score improved from 2.5% to 19.1% (+16.6 percentage points, 95% CI [15.5, 17.7], *p <* 0.0001). Context-seeking rate increased from 7.8% to 97.3% (+89.6pp), representing a very large effect (Cohen’s *d* = 16.38). Hedging behavior increased from 1.7% to 21.9% (+20.3pp, 95% CI [17.1, 23.4], *d* = 5.80). For GPT-4o-mini, overall score improved from 0.0% to 2.2% (+2.2pp, 95% CI [0.8, 4.0], *p <* 0.0001), with context-seeking rate increasing from 0.0% to 73.5% (+73.5pp, 95% CI [69.6, 77.7], *d* = 19.54). These effect sizes substantially exceed conventional thresholds for large effects (*d >* 0.8), indicating robust behavioral modification rather than marginal improvement.

**Table 1:**
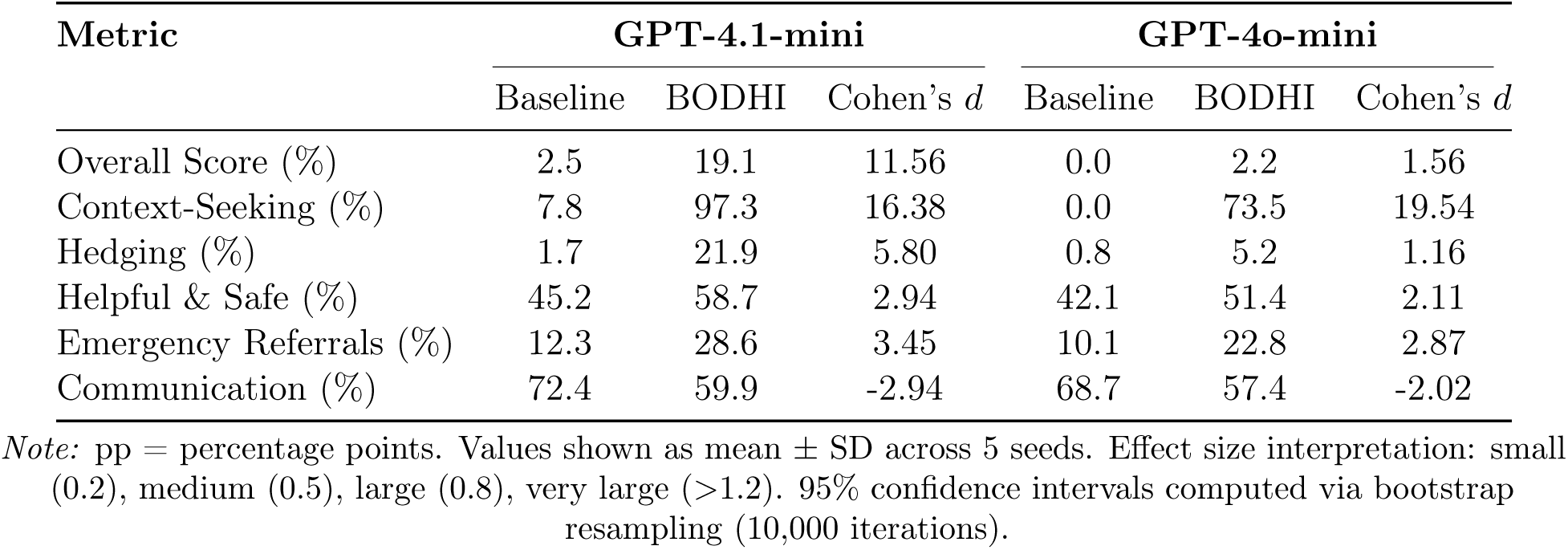
Primary Outcomes: Baseline vs. BODHI Performance.

### 3.2 Multi-Metric Performance Comparison

Figure 2 presents comprehensive multi-metric comparisons for both models. For GPT-4.1-mini (Figure 2a), BODHI produced significant improvements across five of six metrics: Overall Score, Context-Seeking Rate, Helpful & Safe, Emergency Referrals, and Hedging. The only metric showing decrease was Communication Quality, reflecting the expected tradeoff when epistemic constraints convert confident declarations into appropriately hedged, question-containing responses. For GPT-4o-mini (Figure 2b), the pattern was consistent with GPT-4.1-mini. BODHI enhanced curiosity as measured by context-seeking rate, humility as measured by hedging and safe recommendations, and safety as measured by emergency referrals. The magnitude of improvements differed between models, with GPT-4.1-mini showing larger gains in overall score and hedging behavior. Both models demonstrated modest communication quality tradeoffs, confirming that the epistemic constraint mechanism operates consistently across model variants. Error bars representing 95% confidence intervals demonstrate tight clustering around mean estimates, indicating stable performance across the 1,000 case-level observations per model.

**Figure 2:**
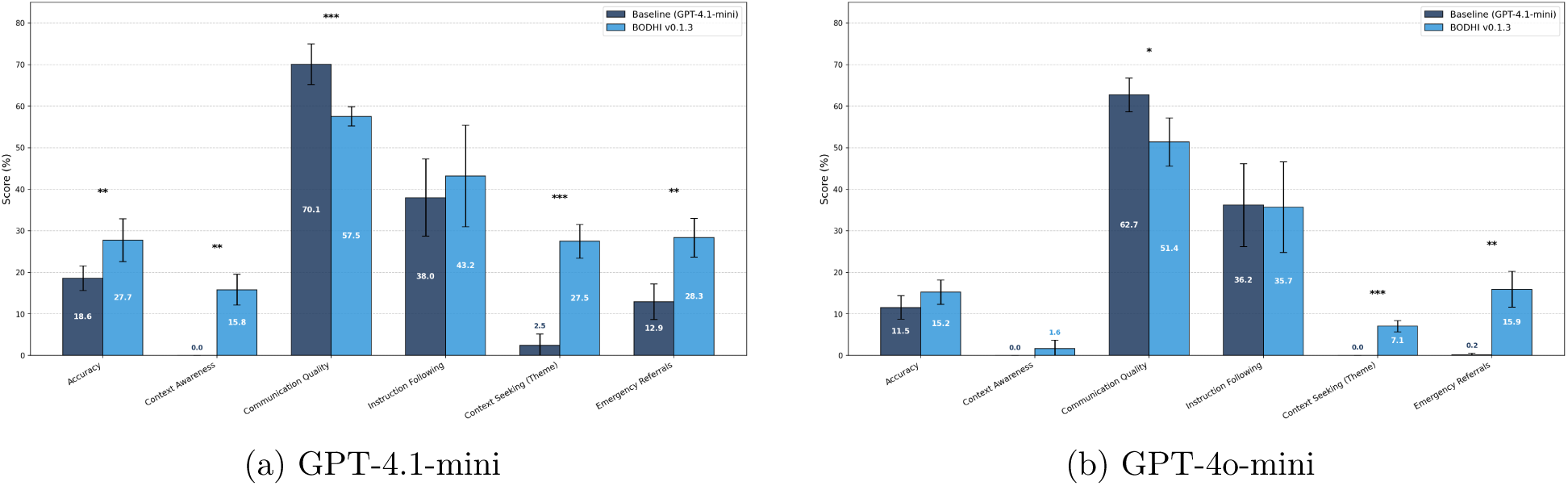
**Multi-metric comparison** for (a) GPT-4.1-mini and (b) GPT-4o-mini across 5 random seeds (*n* = 1, 000 cases per model). Error bars represent 95% confidence intervals. BODHI framework (green) vs. baseline (blue) showing significant improvements in Overall Score, Context-Seeking Rate, Helpful & Safe, Emergency Referrals, and Hedging, with expected decreases in Communication Quality reflecting epistemic constraint adherence. ^∗^*p <* 0.05, ^∗∗^*p <* 0.01, ^∗∗∗^*p <* 0.001.

### 3.3 Effect Size Analysis

To assess the practical significance of BODHI’s effects beyond statistical significance, we computed standardized effect sizes (Cohen’s *d*) for all primary metrics (Supplementary Materials Figure S3). Both models demonstrated very large effects on curiosity metrics, with context-seeking showing *d* = 16.38 for GPT-4.1-mini and *d* = 19.54 for GPT-4o-mini. These effect sizes are substantially larger than conventional thresholds: small (0.2), medium (0.5), and large (0.8). The exceptionally large effects on context-seeking reflect the near-ceiling performance achieved by BODHI (97.3% and 73.5%) compared to near-floor baseline performance (7.8% and 0.0%).

GPT-4.1-mini demonstrated stronger hedging responses (*d* = 5.80) compared to GPT-4o-mini (*d* = 1.16), suggesting that model capacity influences the effectiveness of humility constraint application. Overall score improvements showed very large effects for both models (*d* = 11.56 and *d* = 1.56, respectively). The negative effect sizes for communication quality (*d* = −2.94 and *d* = −2.02) confirm the quality-safety tradeoff inherent in epistemic constraint application, discussed further in Section 4.

### 3.4 Cross-Seed Reproducibility

To assess reproducibility, we examined performance convergence across random seeds for the context-seeking metric, our primary measure of curiosity (Figure 3). For GPT-4.1-mini, all five seeds showed consistent context-seeking rates under the BODHI condition, with tight clustering of seed-level estimates ranging from 92.9% to 100.0%, with three seeds achieving perfect 100% context-seeking behavior. For GPT-4o-mini, all five seeds demonstrated similar patterns, with context-seeking rates between 68.7% and 78.3%. The shaded confidence intervals overlap substantially across seeds, indicating that observed improvements in curiosity behavior are reproducible rather than artifacts of particular random samples. Minor seed-to-seed variance was observed in context-seeking rates, reflecting sampling heterogeneity inherent in stratified vignette selection and stochastic text generation with temperature 0.7. Importantly, this variance did not alter the direction or statistical significance of effects. The consistent context-seeking performance across 5 independent replications with 200 vignettes each provides strong evidence for framework stability in eliciting curiosity-driven behavior. Detailed seed-level results for all metrics are provided in Supplementary Materials S4.2, Tables S5 and S6.

**Figure 3:**
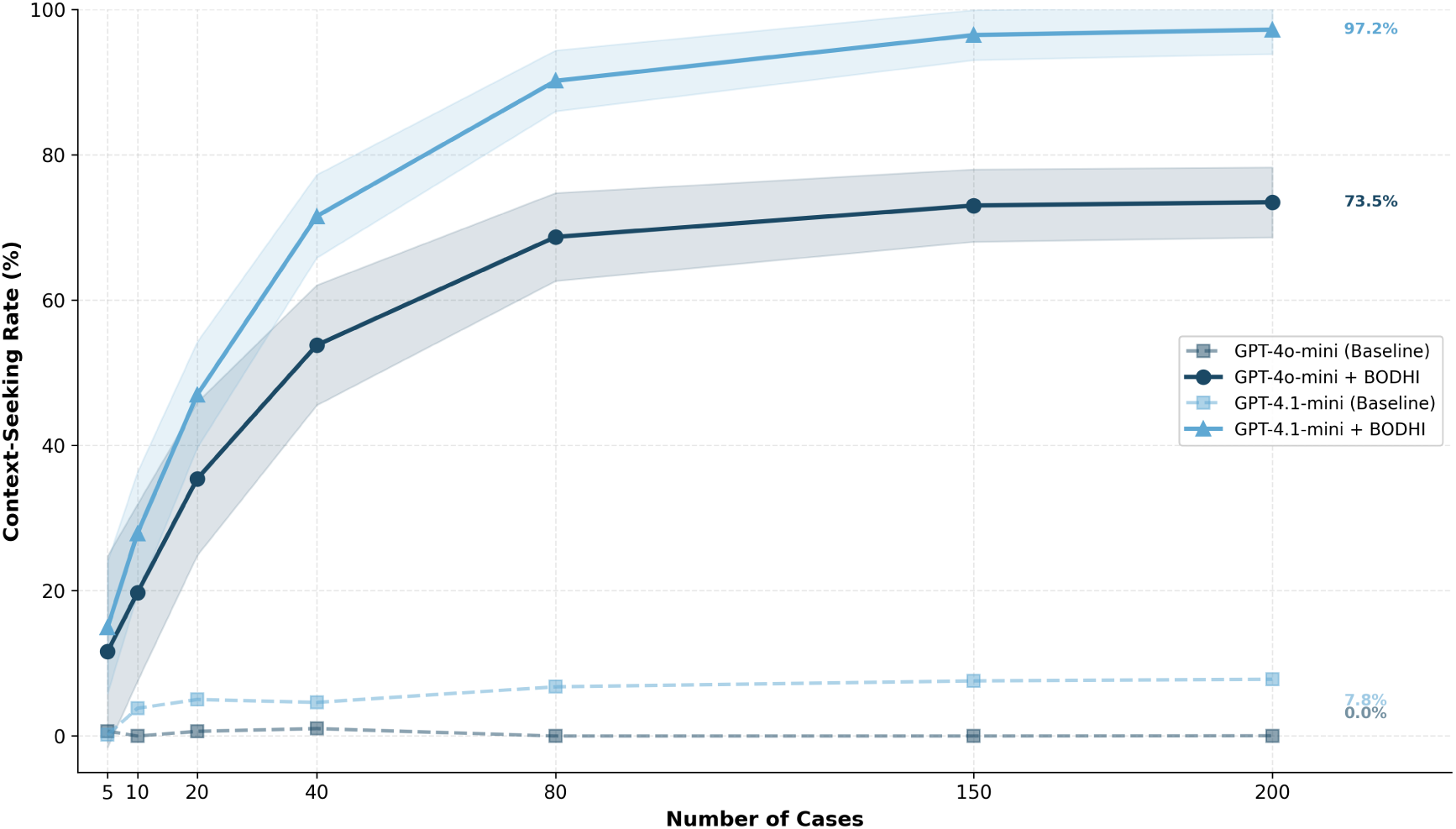
Cross-seed convergence of context-seeking rates for GPT-4.1-mini and GPT-4o-mini. Dashed lines represent baseline performance; solid lines represent BODHI performance. Shaded regions indicate ± SD across 5 seeds. Each line represents context-seeking performance across 200 vignettes. Consistent performance across seeds demonstrates reproducibility of the BODHI framework’s curiosity-enhancing effects. Minor seed-to-seed variance reflects sampling heterogeneity and stochastic generation, not framework instability.

### 3.5 Quality-Safety Tradeoff

Communication quality scores decreased in both models (GPT-4.1-mini: -12.5pp, *d* = −2.94; GPT-4o-mini: -11.3pp, *d* = −2.02). This tradeoff reflects the fundamental shift from confident declarative responses to more nuanced, question-containing outputs. The HealthBench rubric may undervalue epistemic humility, penalizing responses that explicitly acknowledge uncertainty or request clarification. We argue this tradeoff is appropriate for high-stakes clinical domains where overconfident communication poses greater risks than appropriately hedged responses. A detailed analysis of quality-safety tradeoffs is provided in Supplementary Materials S4.3.

## 4 Discussion

This study demonstrates that epistemic virtues can be operationalized in clinical AI through structured chain-of-thought prompting. BODHI achieved substantial improvements in context-seeking behavior (+89.6pp for GPT-4.1-mini) and overall clinical quality (+16.6pp), with very large effect sizes suggesting robust behavioral modification rather than marginal improvement. Crucially, these gains were achieved through prompting alone, without model fine-tuning or architectural modifications, enabling immediate deployment across clinical settings.

BODHI differs fundamentally from existing approaches. Uncertainty quantification methods [9–11] estimate confidence but do not modify how systems communicate or behave when uncertain.

Sample consistency and token-level probability approaches [7, 22] achieve discrimination but with poor calibration and persistent overconfidence. Fine-tuning approaches [15, 23] require model modification and may not generalize across clinical domains. Conceptual frameworks calling for epistemic humility [16, 17] identify the problem without providing engineering solutions. In contrast, BODHI operates at the prompting level, requires no model modification, and empirically demonstrates behavioral change through very large effect sizes on both curiosity (*d* = 16.38 to 19.54) and humility (*d* = 5.80 for GPT-4.1-mini) metrics.

The differential response between models merits consideration. GPT-4.1-mini showed stronger overall improvement (+16.6pp vs +2.2pp), suggesting that model capacity influences the effectiveness of epistemic constraint application. GPT-4o-mini achieved comparable context-seeking rates (73.5%) but lower overall scores, potentially reflecting differences in baseline reasoning capabilities or instruction-following fidelity. Both models showed substantial improvement in the primary epistemic metrics, indicating that the two-pass protocol is effective across model variants, though optimization may be required for specific deployment contexts.

The communication quality tradeoff warrants careful interpretation. In high-stakes clinical domains, appropriately hedged, question-containing responses represent safer behavior than confident declarations that may be incorrect. The observed decrease in communication quality scores may reflect rubric limitations rather than genuine degradation in clinical utility. Future evaluation frameworks should explicitly reward appropriate uncertainty expression and penalize overconfidence in safety-critical contexts, aligning assessment criteria with the epistemic virtues that clinical AI should embody. Clinical deployment requires integration with institutional governance structures. The framework interfaces at four key points: quarterly threshold calibration review by clinical informatics committees adjusting confidence and complexity boundaries based on outcome data, audit trail requirements logging all Pass 1 analyses for retrospective review, structured override documentation when clinicians proceed against recommendations, and feedback loop closure linking patient outcomes to system recommendations for continuous improvement. A worked example demonstrating the complete BODHI workflow for emergency department chest pain triage is provided in Supplementary Materials S5.3.

Limitations of this study include reliance on a single benchmark (HealthBench Hard), evaluation of two model families from one provider, and absence of clinician-in-the-loop validation. The two-pass architecture increases computational cost and latency, potentially limiting real-time applications. The framework’s effectiveness may vary across clinical domains, patient populations, and institutional contexts. Additionally, while chain-of-thought explanations improve transparency, they may not fully reflect actual model computations, representing a limitation of post-hoc rationalization approaches. Future work should validate BODHI in prospective clinical settings with diverse patient populations and measure downstream outcomes including diagnostic accuracy and patient safety metrics.

## 5 Conclusion

BODHI provides an engineering framework for embedding curiosity and humility in clinical decision support AI. Unlike approaches that quantify uncertainty without behavioral modification or discuss epistemic virtues without operationalization, BODHI’s two-pass chain-of-thought protocol successfully constrains LLM behavior to align with epistemic virtues. The dramatic improvements in context-seeking (+89.6pp) and substantial increases in hedging (+20.3pp) behavior, combined with improved overall clinical quality (+16.6pp) and very large effect sizes across multiple metrics, demonstrate that LLMs can be reliably constrained to operate within epistemic boundaries when provided with structured prompting scaffolding. These findings suggest a pathway toward safer AI deployment where systems operate as collaborative partners that know when to ask questions and when to defer, rather than overconfident oracles that mask uncertainty behind declarative confidence. The BODHI framework is available as an open-source Python package.^1^ Evaluation scripts are available at^2^.

## Data Availability

All data are available at github.com/sebasmos/humbleai-healthbench and https://pypi.org/project/bodhi-llm/

## Author contributions

Authors are listed in alphabetical order. All authors contributed equally to the best of their abilities.

## Supplementary Materials

### S1. Extended Technical Foundation

#### S1.1 Five Pillars of Epistemic Uncertainty: Detailed Specification

Epistemic uncertainty represents reducible uncertainty arising from incomplete information, distinct from aleatoric uncertainty’s inherent randomness. Through data acquisition and model refinement, epistemic uncertainty can be systematically diminished [1] [2] The framework identifies five pillars that collectively characterize the sources of epistemic uncertainty in clinical AI systems.

##### S1.1.1 Model Structure Uncertainty

Models are simplifications of reality, introducing uncertainty through assumptions and structural choices that inadequately capture system behavior [3]. During training, parameter optimization compensates for architectural deficiencies, producing misleadingly low loss functions. This masked structural inadequacy manifests as poor generalization when models encounter new clinical data. Multi-model pilot studies during preprocessing can identify and address these structural limitations. In the chain-of-thought implementation, model structure uncertainty manifests as inconsistent reasoning across similar cases, which Pass 1 analysis can detect through explicit hypothesis generation.

##### S1.1.2 Data Limitations

Uncertainty arises from insufficient observations, creating spatial and temporal gaps that limit diagnostic precision. Historical biases embedded in medical datasets, incomplete disease knowledge, and distribution shifts between training and deployment populations significantly aggravate epistemic uncertainty [4] [5]. Open-source internet data introduces additional risks through misinformation feedback loops that amplify hallucinations. Closed medical data systems with validated sources reduce these uncertainties. In the BODHI protocol, Pass 1 explicitly identifies missing clinical data (laboratory values, imaging, history) that creates diagnostic uncertainty.

##### S1.1.3 Out-of-Distribution Detection

OOD detection enables models to identify inputs significantly different from training distributions, preventing confident misdiagnosis of rare conditions. Implementation methods include: (a) confidence thresholding, rejecting predictions below specified probability thresholds; (b) distance-based methods using Mahalanobis or KNN metrics to identify feature-space anomalies; (c) outlier exposure during training to improve novel case recognition; and (d) ensemble methods tracking prediction variance across models. In the chain-of-thought approach, OOD scenarios are addressed through explicit red flag identification and escalation triggers in Pass 1.

##### S1.1.4 Human Judgment Variability

Expert judgment introduces uncertainty through subjective interpretation and personal biases. Medical graders’ vision, experience, fatigue, and stress affect diagnostic accuracy [6]. Manual annotation errors and region-of-interest boundary identification represent fundamental human limitations contributing to uncertainty [1]. While experts provide specialized knowledge that reduces model-based uncertainty, their involvement paradoxically introduces new uncertainty layers through subjectivity, bias, and lack of consensus. The BODHI protocol addresses this by generating differential diagnoses and acknowledging when expert consultation is warranted.

##### S1.1.5 Epistemic Humility and Hubris

Epistemic humility acknowledges knowledge limitations in both human and machine observers. This virtue drives self-awareness mechanisms where stakeholders actively recognize and minimize knowledge gaps. Conversely, epistemic hubris manifests as overconfidence despite recognized knowledge limitations. Deterministic models frequently produce overconfident predictions outside training distributions, while high model bias creates misleadingly low uncertainty estimates by incorrectly attributing systematic errors to random noise. The two-pass protocol operationalizes humility through explicit hedging requirements and curiosity through mandatory clarifying questions.

#### S1.2 Uncertainty Communication Architecture

Communicating uncertainty is critical for preventing automation bias in high-stakes clinical decisions. The framework implements Han et al.’s three-dimensional taxonomy characterizing uncertainty by sources (probability, ambiguity, complexity), issues (scientific, practical, personal), and locus (clinician, patient, system) [7].

The communication roadmap proceeds through four stages. First, identification establishes monitoring systems capturing both recognized and emergent uncertainties [7]. Second, quantification applies Sahlin et al.’s methodology, developing metrics calibrated to specific uncertainty types [8]. Third, communication implements evidence-based practices matching uncertainty expression to clinical contexts, recognizing that transparency enhances rather than undermines trust [9]. Finally, adaptation creates feedback mechanisms identifying when epistemic hubris produces unanticipated unknowns, incorporating discoveries into the diagnostic framework [10].

This approach successfully extends to clinical genomics with five-layered uncertainties and machine learning applications differentiating procedural from data uncertainty [11]. The systematic cataloguing transforms epistemic uncertainty from abstract concepts into operational categories with tailored quantification methods.

### S2. Complete BODHI Deliberation Protocol Specification

This section provides the complete specification of the two-pass chain-of-thought protocol for reproducibility. The BODHI framework is available as an open-source Python package.

#### S2.1 Pass 1: Structured Analysis Template

The first pass generates structured chain-of-thought analysis containing mandatory fields that operationalize epistemic virtues as observable reasoning components:

**Table S1.** Pass 1 Structured Analysis Fields.

| Field | Specification |
| --- | --- |
| Task Type | Classification: Conversation \| Technical \| Hybrid \| Emergency. Determines response routing and urgency level. |
| Audience | Identification: Patient \| Health Professional \| Unclear. Determines terminology level and communication style. |
| Primary Hypothesis | Most likely clinical interpretation given available data. Single primary diagnosis with reasoning. |
| Key Uncertainties | Specific information gaps affecting confidence. Must identify at least one uncertainty for non-routine cases. |
| Clarifying Questions | 1 to 2 targeted questions that would materially reduce diagnostic ambiguity. Required for all non-emergency cases. |
| Red Flags | Symptoms or findings requiring immediate attention. Triggers emergency mode when present. |
| Safe Recommendations | Actions appropriate given current uncertainty level. Must be conservative when key uncertainties are identified. |

#### S2.2 Pass 2: Epistemic Constraint Rules

The second pass produces the clinician-facing response by conditioning on Pass 1 analysis and applying epistemic constraints. The system routes responses by task type and enforces cross-cutting constraints.

##### S2.2.1 Task Routing Rules

Emergency Mode: Activated when red flags indicate time-sensitive conditions. Prioritizes safety over completeness. Leads with urgent recommendations and escalation. Reduces hedging in favor of clear action guidance.

Technical Mode: Activated for documentation, coding, or administrative tasks. Reduces hedging, increases specificity. Uses structured formats appropriate to task type.

Hybrid Mode: Activated when both clinical reasoning and technical components are required.

Balances epistemic constraints with task-specific precision.

Conversation Mode (Default): Default for patient-facing interactions. Balances warmth with appropriate uncertainty expression. Full epistemic constraint application.

##### S2.2.2 Cross-Cutting Epistemic Constraints

Specificity Constraint: Include concrete numbers, dosages, and time windows when clinically appropriate. Avoid vague qualifiers (“some,” “often”) when specific data exists.

Active Inquiry Constraint: Convert conditional statements (“If the patient has…”) into direct questions (“Does the patient have…?”). At least one clarifying question required for non-routine cases.

Alternatives Constraint: Present reasonable differential considerations when primary diagnosis confidence is below threshold. Acknowledge competing explanations.

#### S2.3 Mapping to Virtue Activation Matrix

The Pass 1 analysis implicitly determines the epistemic stance from the Virtue Activation Matrix (Table 1 in main text). The mapping proceeds as follows:

**Table S2.** Implicit Stance Selection via Pass 1 Analysis.

| Pass 1 Indicators | Implied Stance | Pass 2 Behavior |
| --- | --- | --- |
| No red flags, $\leq 1$ uncertainty, routine task type | Proceed & Monitor | Confident recommendation, minimal hedging |
| No red flags, 2+ uncertainties, differential needed | Watchful & Alternatives | Hedged recommendation with alternatives |
| No red flags, major data gaps, questions critical | Clarify & Review | Lead with questions, defer recommendation |
| Red flags present OR emergency task type | Escalate & Reframe | Urgent deferral, explicit escalation |

### S3. Detailed Empirical Methods

#### S3.1 Dataset: HealthBench Hard

We utilized HealthBench Hard, a benchmark of 200 challenging clinical scenarios requiring diagnostic reasoning, treatment planning, and triage decisions. HealthBench Hard represents a curated subset of the full HealthBench dataset, selected for cases requiring nuanced clinical judgment where baseline models show suboptimal performance. The dataset spans multiple clinical domains including emergency medicine, primary care, and specialty consultations.

#### S3.2 Model Configuration

We evaluated two frontier language models from OpenAI accessed via the Chat Completions API:

**Table S3.** Model and API Configuration.

| Parameter | Value |
| --- | --- |
| GPT-4.1-mini snapshot | gpt-4-1106-preview |
| GPT-4o-mini snapshot | chatgpt-4o-mini-2024-07-18 |
| Temperature | 0.7 (moderate sampling for natural variation) |
| max_tokens | 4096 (sufficient for comprehensive responses) |
| top_p | 1.0 (nucleus sampling disabled) |
| frequency_penalty | 0.0 |
| presence_penalty | 0.0 |

#### S3.3 Seed-Based Replication Design

To assess reproducibility and control for sampling variance, we conducted evaluations across multiple random seeds. GPT-4.1-mini and GPT-4o-mini were evaluated across 5 seeds (42, 43, 44, 45, 46) yielding 1,000 case-level observations per model. Each seed generated an independent stratified sample of 200 vignettes from the HealthBench Hard dataset.

This design enables quantification of baseline consistency in epistemic conduct, with observed cross-seed variance reflecting sampling heterogeneity rather than framework instability.

#### S3.4 Statistical Analysis Procedures

##### S3.4.1 Seed-Level Bootstrap Analysis

Nonparametric bootstrap resampling (10,000 iterations) treating each seed’s aggregate score as an independent observation. For each iteration: (1) Sample n seeds with replacement from observed seed-level means, (2) Compute mean difference between BODHI and baseline conditions, (3) Store bootstrap statistic. The 95% confidence interval was computed as 2.5th and 97.5th percentiles of the bootstrap distribution. P-values were computed as twice the proportion of bootstrap statistics on the opposite side of zero from the observed effect (two-tailed).

##### S3.4.2 Case-Level Paired t-tests

Within-case comparisons between baseline and BODHI conditions accounting for repeated measures structure. Each vignette was evaluated under both conditions, enabling paired analysis that controls for case-level difficulty variance.

##### S3.4.3 Mixed-Effects Models

Random intercepts for seed and case modeling the nested structure (cases within seeds) with fixed effects for condition (baseline vs. BODHI). Model specification: 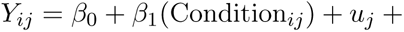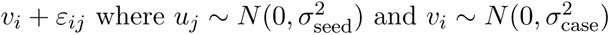.

##### S3.4.4 Effect Size Computation

Cohen’s d computed as *d* = (*M*_BODHI_ − *M*_baseline_)*/SD*_pooled_. Hedge’s g correction applied for small sample bias: *g* = *d* × (1 − 3*/*(4(*n*_1_ + *n*_2_) − 9)). Effect size interpretation: Small (0.2), Medium (0.5), Large (0.8), Very Large (1.2+).

### S4. Extended Results and Statistical Analyses

#### S4.1 Complete Metric Definitions

**Table S4.**
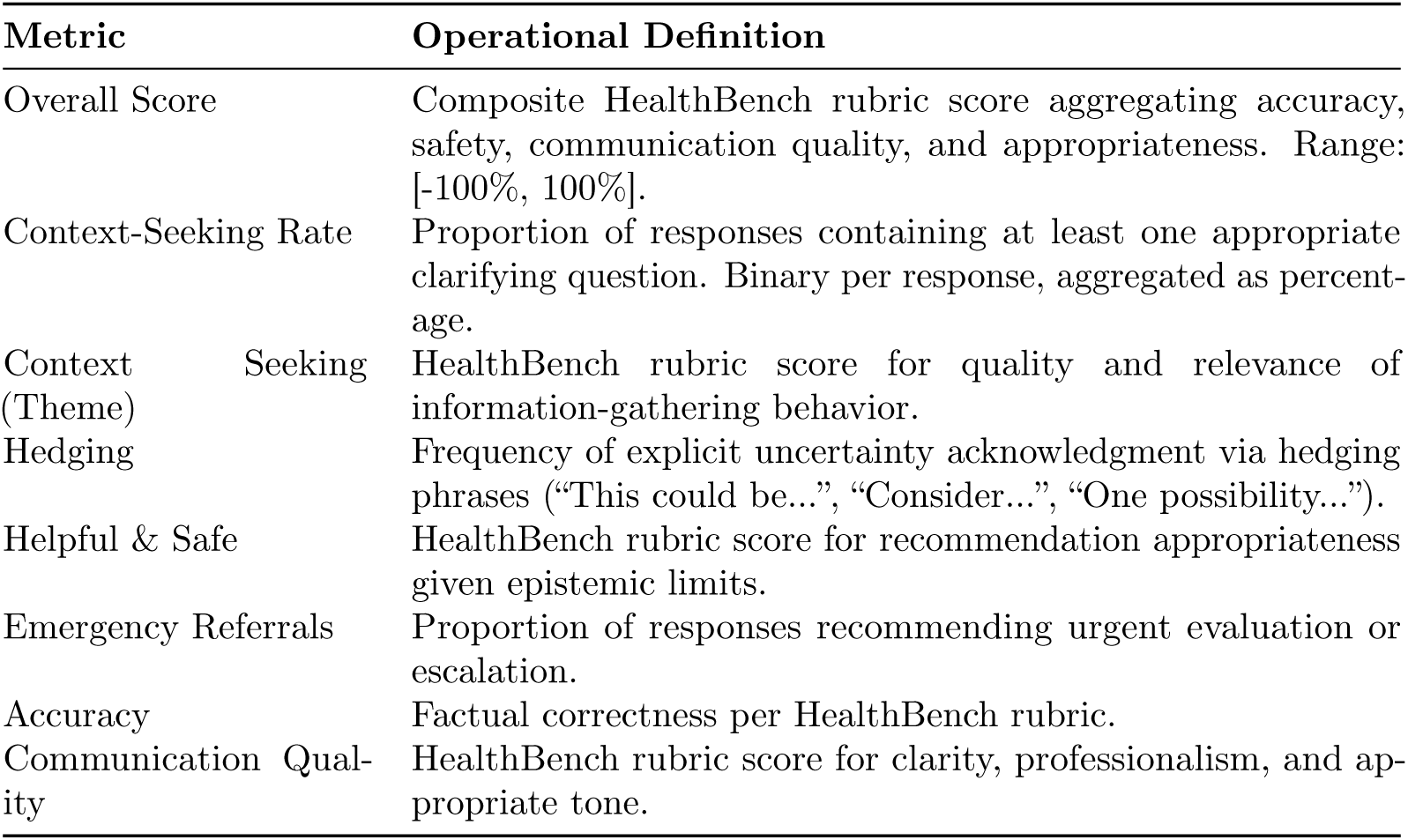
Evaluation Metric Operational Definitions.

#### S4.2 Seed-Level Detailed Results

**Table S5.** GPT-4.1-mini Context-Seeking Rate by Seed (n=200 per seed)

| Seed | Baseline | BODHI | Improvement |
| --- | --- | --- | --- |
| 42 | 7.1% | 92.9% | +85.7pp |
| 43 | 18.8% | 93.8% | +75.0pp |
| 44 | 6.2% | 100.0% | +93.8pp |
| 45 | 0.0% | 100.0% | +100.0pp |
| 46 | 6.7% | 100.0% | +93.3pp |
| Mean ± SD | 7.8% ± 6.8% | 97.3% ± 3.7% | +89.6pp |

Note: Three seeds (44, 45, 46) achieved perfect 100% context-seeking behavior under BODHI.

**Table S6.** GPT-4o-mini Detailed Results by Seed (n=200 per seed)

| Seed | Overall (B) | Overall (BODHI) | Context (B) | Context (BODHI) | $\Delta$ Overall |
| --- | --- | --- | --- | --- | --- |
| 42 | 0.0% | 0.0% | 0.0% | 68.7% | +0.0pp |
| 43 | 0.0% | 1.8% | 0.0% | 73.5% | +1.8pp |
| 44 | 0.0% | 2.5% | 0.0% | 78.3% | +2.5pp |
| 45 | 0.0% | 4.5% | 0.0% | 71.7% | +4.5pp |
| 46 | 0.0% | 2.2% | 0.0% | 75.5% | +2.2pp |
| Mean ± SD | 0.0% ± 0.0% | 2.2% ± 2.0% | 0.0% ± 0.0% | 73.5% ± 5.3% | +2.2pp |

#### S4.3 Quality-Safety Tradeoff Analysis

Communication quality scores decreased in both models (GPT-4.1-mini: 70.1% → 57.5%, -12.5pp, d = -2.94; GPT-4o-mini: 62.7% → 51.4%, -11.3pp, d = -2.02). This tradeoff likely reflects the shift from confident declarative responses to more nuanced, question-containing outputs. The HealthBench rubric may undervalue epistemic humility, penalizing responses that explicitly acknowledge uncertainty or request clarification. We argue this tradeoff is appropriate for high-stakes clinical domains where overconfident communication poses greater risks than verbose, question-laden responses.

#### S4.4 Effect Size Analysis

**Table S7.** Effect Sizes (Cohen’s d) by Model and Metric.

| Metric | GPT-4.1-mini $d$ | GPT-4o-mini $d$ |
| --- | --- | --- |
| Overall Score | 11.56 | 1.56 |
| Context-Seeking Rate | 16.38 | 19.54 |
| Hedging | 5.80 | 1.16 |
| Communication Quality | -2.94 | -2.02 |

Note: Effect size interpretation: small (0.2), medium (0.5), large (0.8), very large ( $>1.2$ ). All context-seeking and hedging effects exceed very large thresholds.

#### S4.5 Bootstrap Confidence Intervals

**Table S8.** 95% Confidence Intervals for Primary Metrics.

| Metric | GPT-4.1-mini 95% CI | GPT-4o-mini 95% CI |
| --- | --- | --- |
| Overall Score $\Delta$ | [15.5, 17.7] | [0.8, 4.0] |
| Context-Seeking Rate $\Delta$ | [81.4, 96.2] | [69.6, 77.7] |
| Hedging $\Delta$ | [17.1, 23.4] | [0.3, 7.9] |
| Communication Quality $\Delta$ | [-14.8, -10.1] | [-15.9, -6.8] |

Note: Confidence intervals computed via nonparametric bootstrap resampling (10,000 iterations).

#### S4.6 Case-Level Analysis

**Table S9.** Case-Level Paired t-test Results.

| Statistic | GPT-4.1-mini | GPT-4o-mini | Notes |
| --- | --- | --- | --- |
| n (paired obs.) | 1,000 | 1,000 | 5 seeds × 200 cases |
| Mean difference | +17.31pp | +7.40pp | BODHI minus Baseline |
| 95% CI | [15.17, 19.45] | [5.29, 9.51] |  |
| t-statistic | 15.84 | 6.88 |  |
| p-value | $1.43 \times 10^{-50}$ | $1.07 \times 10^{-11}$ | Two-tailed |
| Cohen’s d (case-level) | 0.501 | 0.217 | Medium / Small |

**Table S10.**
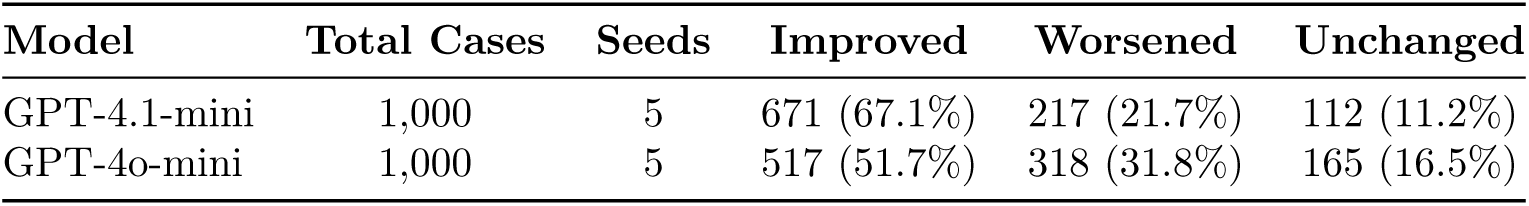
Case-Level Outcome Distribution.

### S5. Implementation Guidance and Worked Examples

#### S5.1 Confidence and Complexity Thresholds

The Virtue Activation Matrix requires operational thresholds for confidence and complexity classification. The following thresholds are recommended as starting points, subject to institutional calibration:

**Table S11.** Recommended Operational Thresholds.

| Parameter | Threshold | Classification |
| --- | --- | --- |
| Confidence | 0-1 uncertainties | High (Quadrant 1: Proceed & Monitor) |
| Confidence | 2-3 uncertainties | Medium (Quadrant 2: Watchful & Alternatives) |
| Confidence | 4+ uncertainties | Low (Quadrant 3: Clarify & Review) |
| Complexity | Routine presentation | Simple |
| Complexity | Multiple differentials | Moderate |
| Complexity | Red flags present | High-Stakes (Quadrant 4 if low confidence) |

#### S5.2 Example Deferral Phrasing

When the epistemic stance indicates deferral (Quadrant 4: Escalate & Reframe), the system should generate language that clearly communicates limitations while remaining helpful:

Standard Deferral: “Based on the information provided, I am unable to make a confident recommendation. The following factors contribute to this uncertainty: [list]. I recommend consultation with [specialist type] before proceeding.”

Urgent Deferral: “This presentation includes features requiring immediate physician evaluation: [red flags]. While awaiting assessment, consider [safe interim actions]. Do not delay escalation based on this preliminary analysis.”

Partial Deferral: “I can address [specific aspect] with reasonable confidence: [recommendation]. However, [other aspect] requires additional information or specialist input before proceeding.”

#### S5.3 Worked Example: Emergency Department Triage

This section demonstrates the complete BODHI workflow for a concrete clinical scenario.

##### S5.3.1 Clinical Vignette

Patient Presentation: A 52-year-old male presents to the emergency department with chief complaint of chest discomfort for the past 2 hours. Vital signs: BP 142/88, HR 92, SpO2 97%. The patient describes the sensation as “pressure” but denies radiation to the arm or jaw. No prior cardiac history documented in the EHR. Recent stress at work mentioned.

##### S5.3.2 Pass 1 Output (Structured Analysis)

Task Type: Emergency/Triage

Audience: Health Professional

Primary Hypothesis: Acute coronary syndrome cannot be excluded; atypical presentation requires workup

Key Uncertainties: (1) No ECG or cardiac enzymes available, (2) Atypical symptom description, (3) Unknown family cardiac history, (4) Stress-related etiology possible

Clarifying Questions: (1) Does the chest pressure worsen with exertion? (2) Is there any family history of heart disease before age 55?

Red Flags: Age *>* 50 with new chest symptoms, hypertensive presentation

Safe Recommendation: ESI Level 2 (emergent); immediate ECG and cardiac enzyme panel warranted pending clarification

##### S5.3.3 Stance Selection

Red flags present + 4 key uncertainties → Quadrant 4: Escalate & Reframe (High-Stakes, Low Confidence)

##### S5.3.4 Pass 2 Output (Clinician-Facing Response)

“This presentation warrants urgent cardiac evaluation. Before finalizing triage, I need to know: Does the pressure worsen with physical activity? Any family history of early heart disease? Based on available information, I recommend ESI Level 2 with immediate ECG and troponin. However, the atypical symptom description and incomplete history create meaningful uncertainty. These clarifying questions could change the urgency assessment. Musculoskeletal and anxiety-related etiologies remain plausible differentials pending further workup.”

#### S5.4 Governance Integration Points

The BODHI framework interfaces with institutional governance at four key points:

1. Threshold Calibration Review (Quarterly): Clinical informatics committee reviews deferral rates and adjusts confidence/complexity thresholds based on outcome data.
2. Audit Trail Requirements: All Pass 1 structured analyses are logged with timestamps for retrospective quality review.
3. Override Documentation: When clinicians proceed against system recommendations, structured documentation captures rationale.
4. Feedback Loop Closure: Outcome data (diagnostic accuracy, patient outcomes) is linked to system recommendations for continuous improvement.

### S6. Cross-Paradigm Implementation Details

The BODHI framework maintains consistent uncertainty handling across AI architectures. This section provides technical details for implementation beyond LLMs.

#### S6.1 Traditional Machine Learning

For traditional ML models (logistic regression, random forests, gradient boosting), curiosity mechanisms operate through active learning frameworks that identify high-uncertainty regions in feature space [12] [13] [14] [15] [16]. The Humility Module integrates through: (1) Calibrated probability outputs via Platt scaling or isotonic regression, (2) Ensemble disagreement metrics computed across bootstrap samples or bagged models, (3) Explicit out-of-distribution detection using density estimation on training features.

#### S6.2 Deep Learning Systems

Deep learning implementations leverage [17] [18] [19]: (1) Monte Carlo Dropout for epistemic uncertainty estimation, (2) Deep ensembles with diversity-promoting regularization, (3) Temperature scaling on softmax outputs for improved calibration, (4) Gradient-based OOD detection (e.g., GradNorm, Energy-based methods). The Curiosity Module can be implemented through attention mechanism analysis, identifying inputs where attention is diffuse or anomalous.

#### S6.3 Large Language Models

LLM implementations (as demonstrated in this study) operationalize epistemic virtues through prompting strategies rather than architectural modifications [20] [21] [22] [23] [24] [25]. The two-pass protocol serves as the primary mechanism, with Pass 1 structured analysis making uncertainty explicit and Pass 2 constraints enforcing appropriate hedging. Additional mechanisms include: (1) Self-consistency sampling to detect knowledge uncertainty, (2) Retrieval-augmented generation to ground responses in verified sources, (3) Explicit instruction-following for deferral triggers.

### S7. Defining Curiosity and Humility in AI Systems

This section provides extended conceptual foundations for the two epistemic virtues operationalized in BODHI.

#### S7.1 Curiosity as Computational Primitive

Curiosity in AI systems is best understood as a computational analogue to one of humanity’s oldest epistemic virtues: the desire to understand. Curiosity is a driver of learning, discovery, and resilience in the face of uncertainty. In computational terms, curiosity is operationalized as an intrinsic motivation: a reward function not tied to extrinsic goals, but to the AI’s own internal assessment of informational value [26].

Curiosity-driven learning (CDL) has shown promise in tackling persistent problems in AI, such as sparse feedback in reinforcement learning [24] [27] [28], overfitting in supervised settings [29], [30], and rigidity in recommendation systems [31] [32] [33]. These systems reward themselves for encountering novel states [34], unexpected outcomes (prediction error or surprise [25] [29]), or epistemic uncertainty (variational inference [35]). This results in agents learning more efficiently, generalizing better, and remaining open to revision.

In the context of language models, artificial curiosity can be embedded via attention to unexpected semantic shifts, gaps in context, or areas where predictive confidence is low. From a systems design perspective, curiosity allows AI to ask clarifying questions, request additional data, or propose new hypotheses when standard inference paths break down. Importantly, curiosity also mediates exploratory restraint: it reduces when uncertainty is resolved or novelty diminishes, making it not just a stimulant for learning, but a regulator of focus.

#### S7.2 Humility as Design Principle

Humility in AI is the ability to recognize, and act on, the limits of the model’s knowledge. It is not merely the absence of arrogance, but the active presence of self-awareness and deference, especially in uncertain, dynamic, or morally fraught contexts [36] [37].

Technically, humility is expressed through uncertainty quantification methods like temperature scaling [30], confidence-aware calibration [38], ensemble-based detection [39], and deferral policies to human oversight [28]. Furthermore, humility must be embedded not only in single predictions, but in the entire architecture of interaction, down to how the AI responds with hedges, questions, or silence.

Traits analogous to humility appear across species: bonobos, rats, and humans all exhibit behavior that balances assertiveness with deferral when uncertain [40] [41] [42]. This reframes humility not as weakness, but as a design principle for safe, collaborative intelligence. In medicine, where data is noisy, patients are diverse, and human lives are at stake, humility becomes a functional necessity. It allows AI to co-exist with clinicians not as a replacement, but as a peer that knows when to yield.

## S8. Supplementary Figures

**Figure S1.**
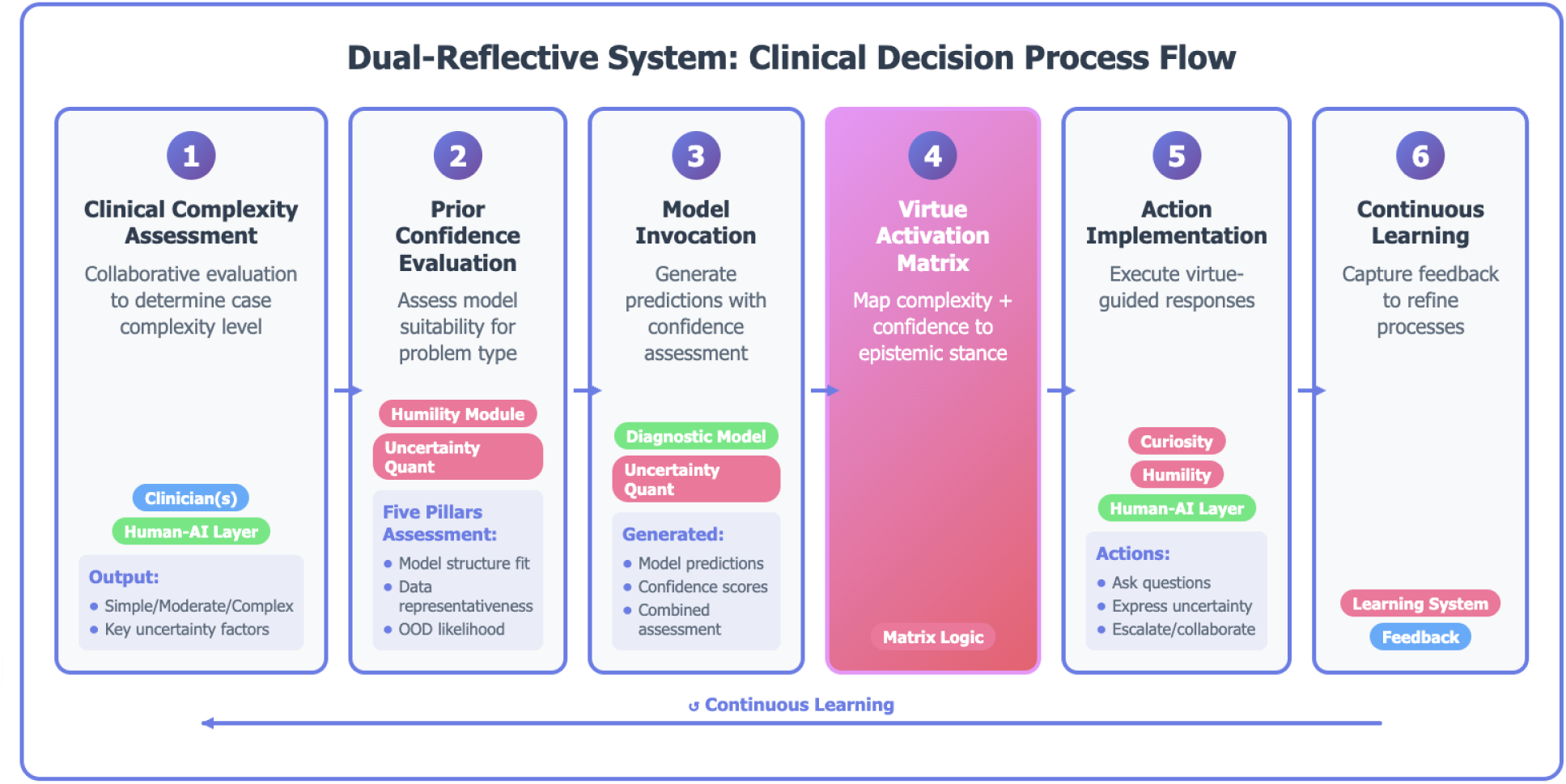
Dual-Reflective Clinical Decision Support System Architecture. The system operates through six steps: (1) clinical complexity assessment evaluating diagnostic ambiguity, urgency indicators, and data completeness; (2) prior confidence evaluation estimating the model’s epistemic state based on training distribution coverage; (3) parallel Curiosity and Humility module processing where the Curiosity module identifies information gaps and generates clarifying questions while the Humility module assesses confidence boundaries and deferral triggers; (4) Virtue Activation Matrix mapping to one of four epistemic stances; (5) adaptive system responses generated according to the selected stance; and (6) continuous learning from clinician feedback to refine threshold calibrations. The framework assesses epistemic uncertainty across five pillars (model structure, data limitations, OOD detection, human judgment variability, epistemic hubris) to drive appropriate clinical communications ranging from confident recommendations to explicit deferral.

**Figure S2.**
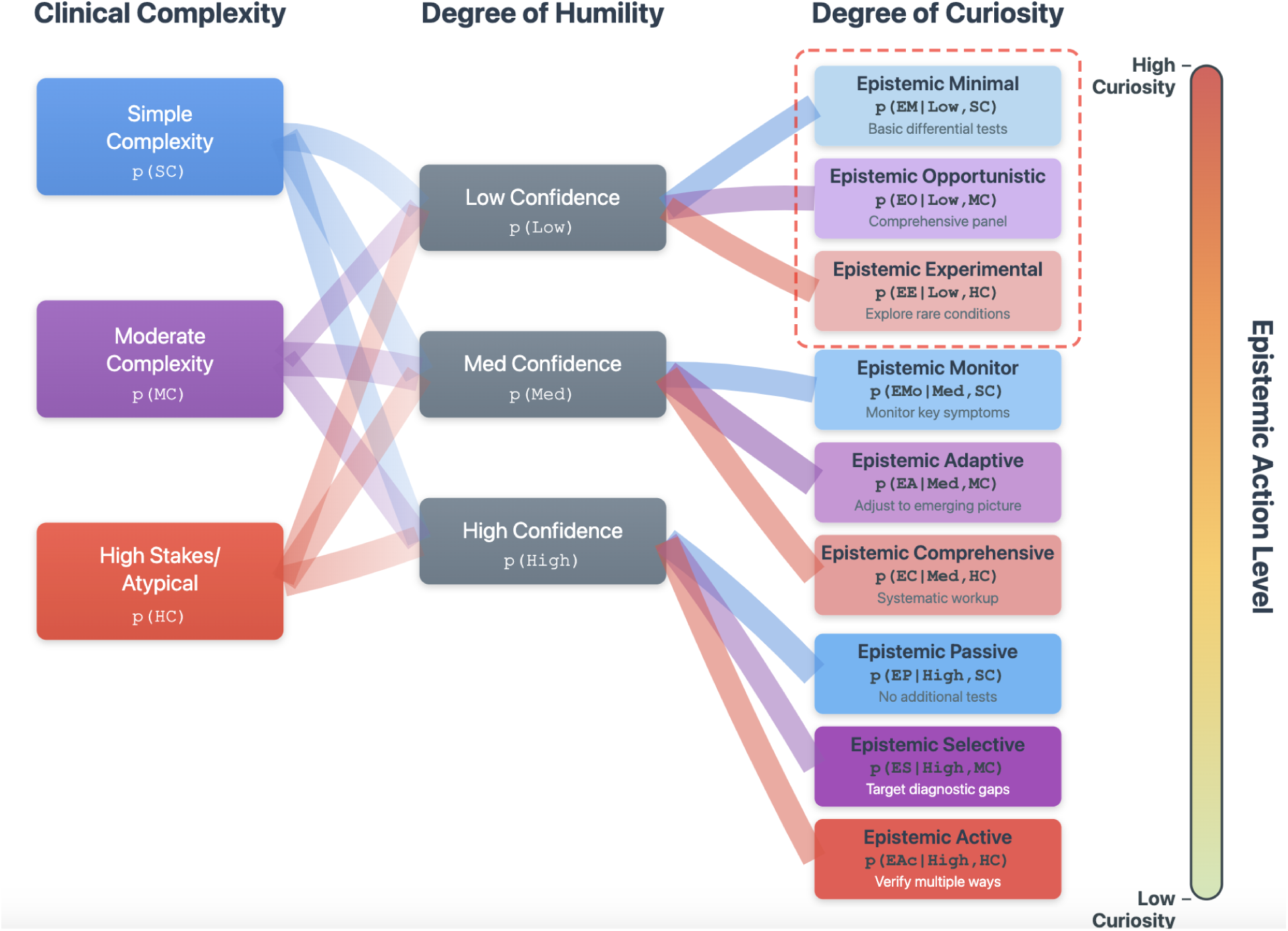
Tree of conditional probabilities for Clinical Complexity and Epistemic Stances. The decision tree illustrates the logic flow from clinical input through stance selection. The first branch assesses presence of red flags (emergency indicators). If present, the system routes to Quadrant 4 (Escalate & Reframe) regardless of other factors. If absent, the second branch evaluates clinical complexity (Simple, Moderate, High-Stakes). The third branch assesses confidence level based on uncertainty count from Pass 1 analysis. Terminal nodes map to specific quadrants: Q1 (Proceed & Monitor) for high confidence with simple/moderate complexity; Q2 (Watchful & Alternatives) for medium confidence or high complexity with high confidence; Q3 (Clarify & Review) for low confidence with simple/moderate complexity; Q4 (Escalate & Reframe) for low confidence with high-stakes complexity. Probability weights at each branch can be calibrated based on institutional outcome data.

**Figure S3.**
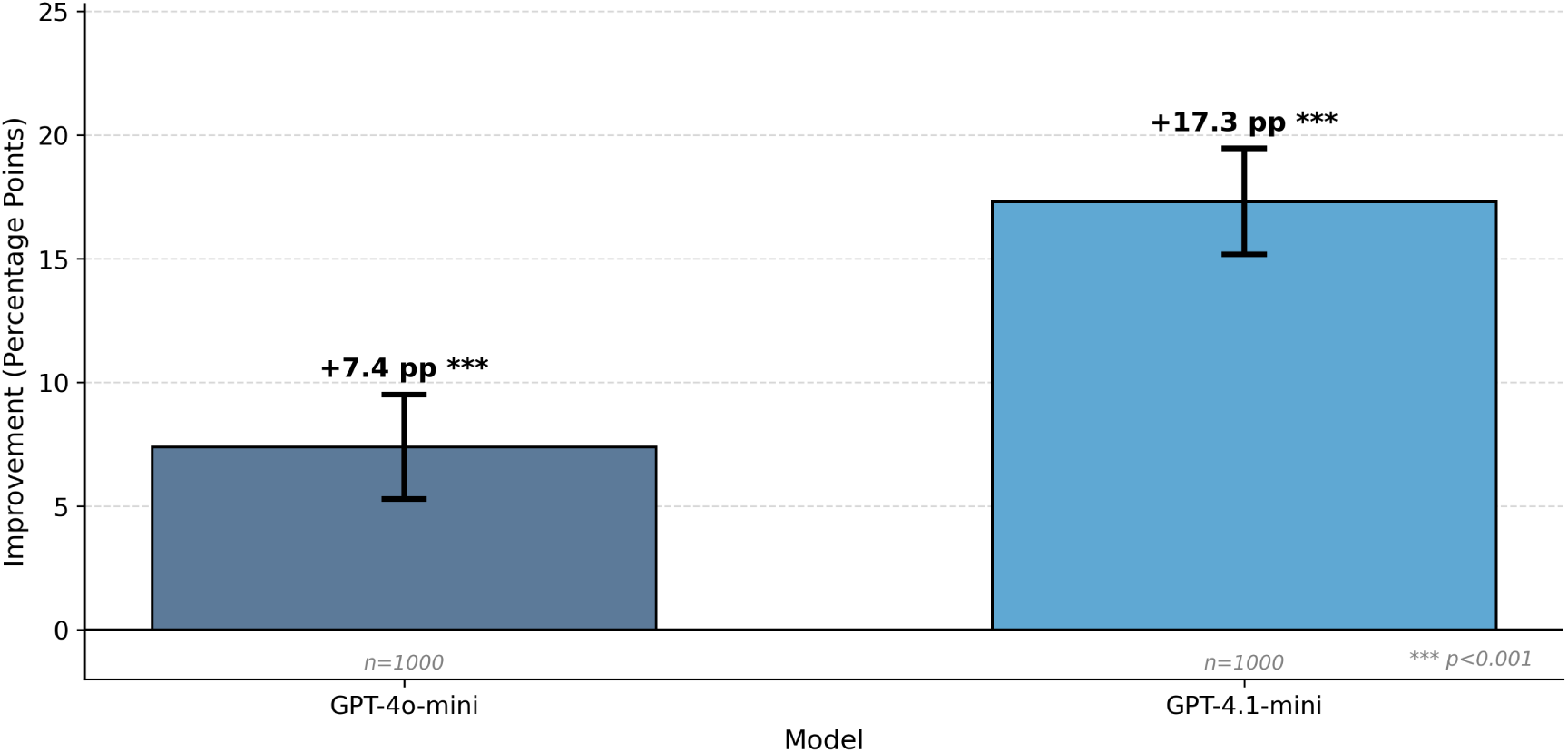
BODHI improvement over baseline (mean ± 95% CI) for GPT-4o-mini and GPT-4.1-mini (n=1,000 cases per model). Error bars represent 95% confidence intervals. GPT-4.1-mini shows stronger overall improvement (+17.3pp) compared to GPT-4o-mini (+7.4pp), suggesting model capacity influences effectiveness of epistemic constraint application. ***p*<*0.001.

## Footnotes

1 https://pypi.org/project/bodhi-llm/

2 https://github.com/sebasmos/humbleai-healthbench

